# India Hypertension Control Initiative- Blood Pressure Control using Drug and Dose-Specific Standard Treatment Protocol at Scale in Punjab and Maharashtra, India, 2022

**DOI:** 10.1101/2023.08.17.23294195

**Authors:** Prabhdeep Kaur, Manikandanesan Sakthivel, Vettrichelvan Venkatasamy, Padmaja Jogewar, Sandeep S Gill, Abhishek Kunwar, Meenakshi Sharma, Anupam Khungar Pathni, Kiran Durgad, Swagata Kumar Sahoo, Amol Wankhede, Navneet Kumar, Vishwajit Bharadwaj, Bidisha Das, Tejpalsinh Chavan, Suhas Khedkar, Lalit Sarode, Sampada D Bangar, Ashish Krishna, Roopa Shivashankar, Parasuraman Ganeshkumar, Pragati Pragya, Balram Bhargava

## Abstract

**Background:** Hypertension treatment coverage is low in India. A stepwise simple treatment protocol is one of the strategies to improve hypertension treatment in primary care. We estimated the effectiveness of various protocol steps to achieve blood pressure (BP) control in public sector health facilities in Punjab and Maharashtra, India, where the India Hypertension Control Initiative (IHCI) was implemented.

**Methods:** We analyzed the records of people enrolled for hypertension treatment and follow-up under IHCI between January 2018 and December 2021 in public sector primary and secondary care facilities across 23 districts from two states. Each state followed a different treatment protocol. We calculated the proportion with controlled BP at each step of the protocol. We also estimated the mean decline in BP pre-and post-treatment.

**Results:** Of 281,209 patients initiated on Amlodipine 5 mg, 159,292 continued on protocol drugs and came for a follow-up visit during the first quarter of 2022. Of 33,450 individuals who came for the follow-up in Punjab and 125,842 in Maharashtra, 70% and 76% had controlled BP, respectively, at the first step with Amlodipine 5 mg. In Punjab, at the second step with amlodipine 10 mg, the cumulative BP control increased to 75%. A similar 5% (76% to 81%) increase was seen in the second step after adding Telmisartan 40 mg in Maharashtra. Overall, the mean (SD) systolic blood pressure (SBP) decreased by 16 mmHg from 148 (15) mmHg at the baseline in Punjab. In Maharashtra, the decline in the mean (SD) SBP was about 15 mmHg from the 144 (18) mmHg baseline.

**Conclusion:** Simple drug- and dose-specific protocols helped achieve a high control rate among patients retained in care under program conditions. We recommend treatment protocols starting with a single low-cost drug and escalating with the same or another antihypertensive drug depending on the cost and availability.

## Introduction

In India, an estimated 28.5% of adults have hypertension; of people with hypertension, 14.5% were on treatment in 2018-19 [1]. Treatment of hypertension is a cost-effective intervention to reduce cardiovascular mortality and morbidity. As per the global monitoring framework for monitoring noncommunicable diseases (NCD), one indicator is a 25% relative reduction in raised blood pressure [2]. Rapid scale-up of diagnosis and treatment coverage is necessary to achieve this goal. The management of hypertension requires a combination of lifestyle modification and treatment with anti-hypertensives. Various guidelines provide detailed information about best practices for managing hypertension and are used by physicians to make informed decisions [3,4].

In India, the National Programme for Prevention & Control of Non-Communicable Diseases (NP-NCD) is a national government program to improve screening and treatment for NCDs. Including hypertension [5]. In 2017, India Hypertension Control Initiative (IHCI) was launched to strengthen the hypertension component of NP-NCD in primary care public sector facilities in India [6]. The initiative aimed to build the capacity of all levels of healthcare workers to diagnose and treat hypertension. We developed simple, stepwise treatment protocols to train doctors and to ensure the availability of a few low-cost anti-hypertensive drugs in primary care health facilities. A treatment protocol is a simplified version of a guideline and is drug and dose-specific. The IHCI team collaborated with state NCD teams, experts, and program managers to conduct consensus meetings to develop and establish a protocol in each state. The program managers from the state health department, physicians from the district hospitals, specialists working in medical colleges, and external experts attended the consensus meetings. Since 2018, 26 states/union territories have developed and implemented treatment protocols. The protocols include three drugs: Amlodipine, Telmisartan (except in one state, which selected Enalapril), and one of the diuretics, chlorthalidone or hydrochlorothiazide [7]. All protocols start with Amlodipine 5 mg for people diagnosed with hypertension except those with chronic kidney disease, for whom Telmisartan was recommended. In the second step, either Amlodipine is escalated to 10 mg or Telmisartan 40 mg is added, depending on the protocol the state chose. IHCI was initially implemented in 26 districts across five states, and early outcomes have been published elsewhere [6].

Prescription practices have aligned with the protocols over time among people newly diagnosed with hypertension [8]. However, until now, we have not analyzed how many people who initiated treatment were controlled at various protocol steps in the project sites. This information is important in optimizing the protocols for scaling treatment in the program setting. Hence, we estimated the effectiveness of various protocol steps to achieve blood pressure control in public sector health facilities in Punjab and Maharashtra, India. We also analyzed the decline in systolic blood pressure (SBP) and diastolic blood pressure (DBP) among people treated according to protocols.

## Methods

### Study design, population, and setting

We analyzed the treatment records of people who were enrolled for hypertension treatment and follow-up as per treatment protocol under IHCI between January 2018 and December 2021 across 23 districts of two states in India, namely Maharashtra and Punjab. We included people with hypertension already treated with Amlodipine 5mg or newly initiated on Amlodipine 5 mg as per the protocol in these two states. We chose these two states for our study as the IHCI was first initiated in these states, and a digital database was available for in-depth analysis. Since the two states used different treatment protocols, we could compare blood pressure control at various steps. We tracked the cohort of people on treatment at primary and secondary public sector health facilities in these IHCI districts. The primary care facilities included health and wellness centers (HWC) and primary health centers (PHC). The secondary care facilities included community health centers (CHCs), sub-district hospitals (SDH), and district hospitals (DH). We have provided detailed descriptions of various types of facilities elsewhere [6].

### Description of the intervention

The IHCI project strengthened hypertension treatment by introducing simple drug and dose-specific treatment protocols adopted in consultation with various stakeholders at the state level [7,9]. The project developed the capacity of healthcare workers at all levels, streamlined the drug supply chain and promoted patient-centric approaches with 30-day drug refills at all levels of health facilities. The project has been described in detail elsewhere [6].

Registrations under IHCI included the patients newly diagnosed with hypertension and those with known hypertension. At the time of registration, the patient’s demographic information, blood pressure, diagnosis, and medications were recorded in the mobile-based digital information system, namely the “Simple” app [10]. The nurses could register a new patient within 80 seconds and document a follow-up visit within 15 seconds using the “Simple” app. The nurses at the health facilities documented patients’ registrations and follow-up visits in the “Simple” app using a unique identification number linked to a QR code for each patient available in the BP passports given to the patients [11]. The doctors and nurses could retrieve the patient record within five seconds by scanning the QR code and viewing the previous blood pressure recording and medications. The “Simple” app improved the documentation of blood pressure values during each visit. Also, the app generated the list of overdue patients, with the high-risk patients prioritized at the top. With a single click, the nurses could call overdue patients using a toll-free, anonymized call through the “Simple” app. The patients who missed the visits also received auto-generated reminder messages. The “Simple” app was an open-source tool that enabled real-time blood pressure and medication documentation during patient visits, monitoring progress and performance. The app also automatically generated reports, saving health workers time compiling and verifying paper-based records.

### Data extraction

We used data from the “Simple” app for the current analysis. We retrieved the deidentified data of people on treatment, which included the following variables - district, treatment facility, the date of registration and last two follow-up visits, blood pressure values, and anti-hypertensive drugs taken during registration and follow-up.

### Operational definition

#### Hypertension diagnosis and control

We defined hypertension as systolic blood pressure ≥ 140 mmHg or diastolic blood pressure ≥ 90 mmHg on at least two measurements or if the individual was already on treatment for hypertension, irrespective of the blood pressure. We graded hypertension as Grade 1 – SBP 140 to 159 mmHg or DBP 90 to 99 mmHg, Grade 2 more than or equal to SBP 160, or DBP more than or equal to 100 mmHg [4]. We considered an individual to have controlled blood pressure (BP) if their SBP was less than 140 mmHg and DBP was less than 90 mmHg [4].

We considered a registered patient as lost-to-follow-up if the patient had not visited the health facility for hypertension treatment even once between January 2022 and March 2022.

#### Treatment protocols

Since the IHCI treatment protocols were tailored to each state in consultation with the state-level stakeholders, the two states included in the current analysis had different hypertension treatment protocols [7].

In Punjab, all individuals diagnosed with hypertension were started on amlodipine 5 mg at the registration facility by the medical officers and reviewed after 30 days (Supplementary Figure 1). For those with uncontrolled BP, the dose of amlodipine was increased to 10 mg and reviewed every 30 days. If the individual had uncontrolled BP during the review, telmisartan 40 mg was added and further escalated to 80 mg if required. Chlorthalidone 12.5 mg was added for those with uncontrolled BP and further escalated to Chlorthalidone 25 mg.

In Maharashtra, all individuals diagnosed with hypertension were started on amlodipine 5 mg and reviewed every 30 days (Supplementary Figure 2). However, if the BP was uncontrolled, the following two treatment escalation steps included the addition of 40 mg and 80 mg of telmisartan. The further escalation steps were increasing the dose of amlodipine to 10 mg and adding Chlorthalidone 6.25 mg and 12.5 mg.

### Data analysis

We extracted the data from the “Simple” app to an Excel spreadsheet and analyzed using STATA software version 17.0 [12]. Among the individuals registered with hypertension under IHCI in the two states between January 2018 and December 2021, we included those who were only on amlodipine 5 mg during registration for the analysis. We excluded those who were lost-to-follow-up and were on non-IHCI protocol drugs at follow-up. We described the individuals included and excluded in the final analysis by age group, gender, presence of diabetes mellitus, type of facility, and grades of hypertension using proportion. We calculated the mean (SD) of systolic and diastolic blood pressure during registration (baseline) and the duration of follow-up. We compared the mean (SD) systolic and diastolic blood pressure between baseline and follow-up using paired t-test by different categories of age, gender, diabetes mellitus, and facility type. We excluded those individuals with missing BP values during the follow-up for this analysis. We considered a p-value less than 0.05 as statistically significant. We calculated the proportion and cumulative proportion of individuals with controlled BP at each step of the IHCI treatment protocol based on the combination of drugs the patient was taking during the second last visit. We did this analysis by different grades of hypertension at the baseline. We conducted and presented the analysis separately for each state, as the treatment protocols differed.

### Human subject protection

We used anonymized records of patients registered under IHCI for the current analysis. The IHCI project was approved by the Institute Human Ethics Committee (IHEC) of the Indian Council of Medical Research – National Institute of Epidemiology (ICMR-NIE), Chennai, with the reference number NIE/IHEC/201709-02.

## Results

Between January 2018 and December 2021, 281,209 individuals were enrolled under the IHCI project in the two states and treated with Amlodipine 5 mg as the first-line drug. Of these, we excluded 100,515 (36%) individuals who did not return to care between January 2022 to March 2022 and 21,402 (8%) who were not on any of the IHCI protocol regimens at follow-up for the analysis. Hence, the final analysis included 159,292 individuals with hypertension, with 79% (125,842) being from Maharashtra (Figure 1).

**Figure 1.**
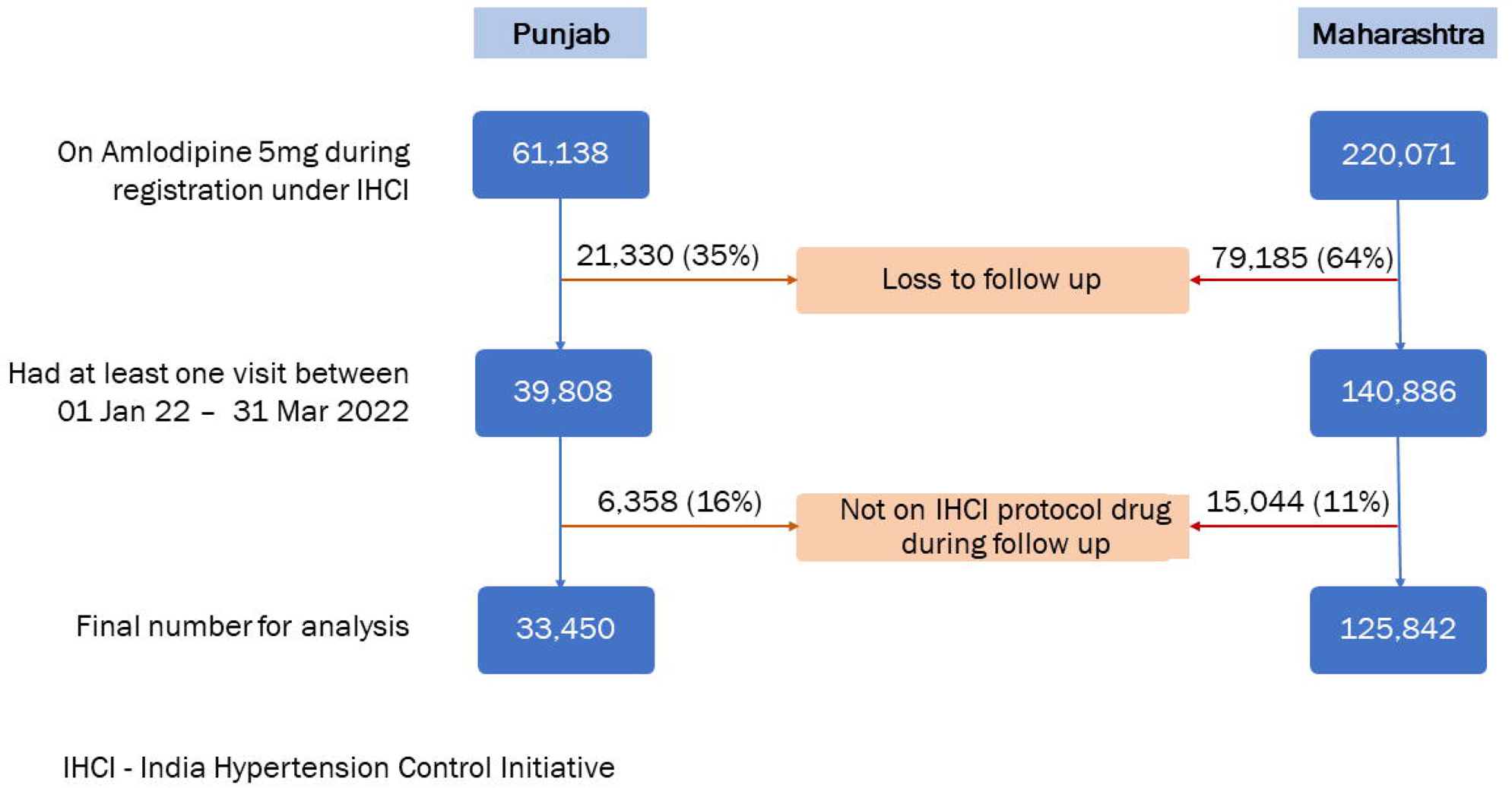
Inclusion and exclusion of individuals with hypertension enrolled under IHCI between January 2018 and December 2022 for the analysis

Of the 159,292 individuals, 95,620 (60%) were 50-69 years and 100,532 (63%) were females (Table 1). There were 12,038 (8%) patients with diagnosed diabetes mellitus, ranging from 5% in Maharashtra to 16% in Punjab. About 86% of the individuals were registered in primary health care facilities (Primary Health Centres or Health and Wellness Centres). This proportion was higher in Maharashtra (88%) when compared with Punjab (80%). About 45,151 (28%) individuals had controlled blood pressure at the time of registration [Punjab vs. Maharashtra – 17% vs. 32%], 73,152 (46%) had Grade I hypertension [Punjab vs Maharashtra – 56% vs. 43%], and 40,989 (26%) had Grade II hypertension [Punjab vs. Maharashtra – 28% vs 25%]. BP was not recorded for about 485 (1%) in Punjab and 2,645 (2%) in Maharashtra during registration. The median (IQR) duration of follow-up was 12 (7-20) months, with Punjab [12 (7-22) months] and Maharashtra [13 (7-20)] having similar follow-up duration.

**Table 1.** Characteristics of individuals with hypertension registered under IHCI from Jan 2018 to Dec 2021 and had at least one follow-up visit between 01 Jan 2022 to 31 Mar 2022, included* in the analysis.

| Characteristic | Category | Punjab (N=33,450) |  | Maharashtra (N=125,842) |  | Total (N=159,292 ) |  |
| --- | --- | --- | --- | --- | --- | --- | --- |
|  |  | n | % | n | % | n | % |
| Age group (in years) | <30 | 145 | 0.4 | 247 | 0.2 | 392 | 0.2 |
|  | 30-49 | 7,154 | 21.4 | 18,361 | 14.6 | 25,515 | 16.0 |
|  | 50-69 | 19,606 | 58.6 | 76,014 | 60.4 | 95,620 | 60.0 |
|  | ≥70 | 6,545 | 19.5 | 31,220 | 24.8 | 37,765 | 23.7 |
| Gender | Male | 10,266 | 30.7 | 48,494 | 38.5 | 58,760 | 36.9 |
|  | Female | 23,184 | 69.3 | 77,348 | 61.5 | 1,00,532 | 63.1 |
| Diabetes | Yes | 5,308 | 15.9 | 6,730 | 5.4 | 12,038 | 7.6 |
|  | No | 28,142 | 84.1 | 1,19,112 | 94.6 | 1,47,254 | 92.4 |
| Facility type | SDH/ DH | 1,993 | 6.0 | 11,154 | 8.9 | 13,147 | 8.3 |
|  | CHC | 4,647 | 13.9 | 4,464 | 3.5 | 9,111 | 5.7 |
|  | PHC | 4,392 | 13.1 | 48,620 | 38.6 | 53,012 | 33.3 |
|  | HWC | 22,418 | 67.0 | 61,604 | 48.9 | 84,022 | 52.7 |
| Baseline BP grades | Controlled BP | 5,524 | 16.5 | 39,627 | 31.5 | 45,151 | 28.3 |
|  | Grade I | 18,599 | 55.6 | 54,553 | 43.3 | 73,152 | 45.9 |
|  | Grade II | 9,327 | 27.8 | 31,662 | 25.1 | 40,989 | 25.7 |
| Baseline SBP (mmHg) | Mean (SD) | 147.8 (15.0) |  | 143.8 (17.7) |  | 144.7 (17.3) |  |
| Baseline DBP (mmHg) | Mean (SD) | 88.0 (9.5) |  | 86.5 (11.0) |  | 86.9 (10.7) |  |
| Treatment duration in months | Median (IQR) | 12 (7 - 22) |  | 13 (7 - 20) |  | 12 (7-20) |  |

\*Those registered under IHCI up to Dec 2021 and taking only Amlodipine 5mg during registration and had at least one follow-up visit between 01 Jan 2022 to 31 Mar 2022 and on any of the drug regimens as per IHCI protocol during the recent follow-up
DH – District Hospital; SDH – Sub-district Hospital; CHC – Community Health Centre; PHC – Primary Health Centre; HWC – Health and Wellness Centre

Among the 121,917 individuals excluded from the analysis, 72,771 (60%) belonged to the 50-69 years age group, and 2,07,920 (57%) were females (Supplementary Table 1). The proportion with diabetes (32%) was higher than those included in the analysis. While 76% of the individuals excluded were registered at primary health care facilities (primary health centers or health and wellness centers), 18% were registered at district or sub-district hospitals. About 32% of the excluded individuals had controlled blood pressure at registration, and about 36% had Grade I hypertension. BP was not recorded for about 9% during registration. The individuals who were excluded from the analysis differed from those included in terms of the gender distribution (males – 43% vs. 37%), the proportion with diabetes (32% vs. 8%), and the proportion with controlled BP (32% vs. 28%) and Grade I hypertension (36% vs. 46%) during registration.

The BP values during the recent follow-up visit were missing for 1% (485/33450) individuals in Punjab and 2% (2645/125842) in Maharashtra and were excluded while comparing the BP at baseline and follow-up. In Punjab, the mean (SD) systolic blood pressure (SBP) decreased by 16.2 mmHg from 147.8 (15.0) mmHg at the baseline to 131.7 (11.9) mmHg during the most recent visit (p-value <0.001) (Table 2). In Maharashtra, the decline in the mean (SD) SBP was about 15 mmHg from the baseline [143.8 (17.7) mmHg] to the most recent visit [129.0 (12.1) mmHg] (p-value <0.001). The SBP decline was consistent across all the age groups at follow-up compared to baseline in both states. We observed a similar decline in sub-groups based on gender and diabetes mellitus in Punjab (16-18 mmHg) and Maharashtra (14-16 mmHg). There was a reduction in the mean SBP across all types of health facilities ranging from 16 to 18 mmHg in Punjab and 12 to 15 mmHg in Maharashtra (p-value <0.001).

**Table 2:**
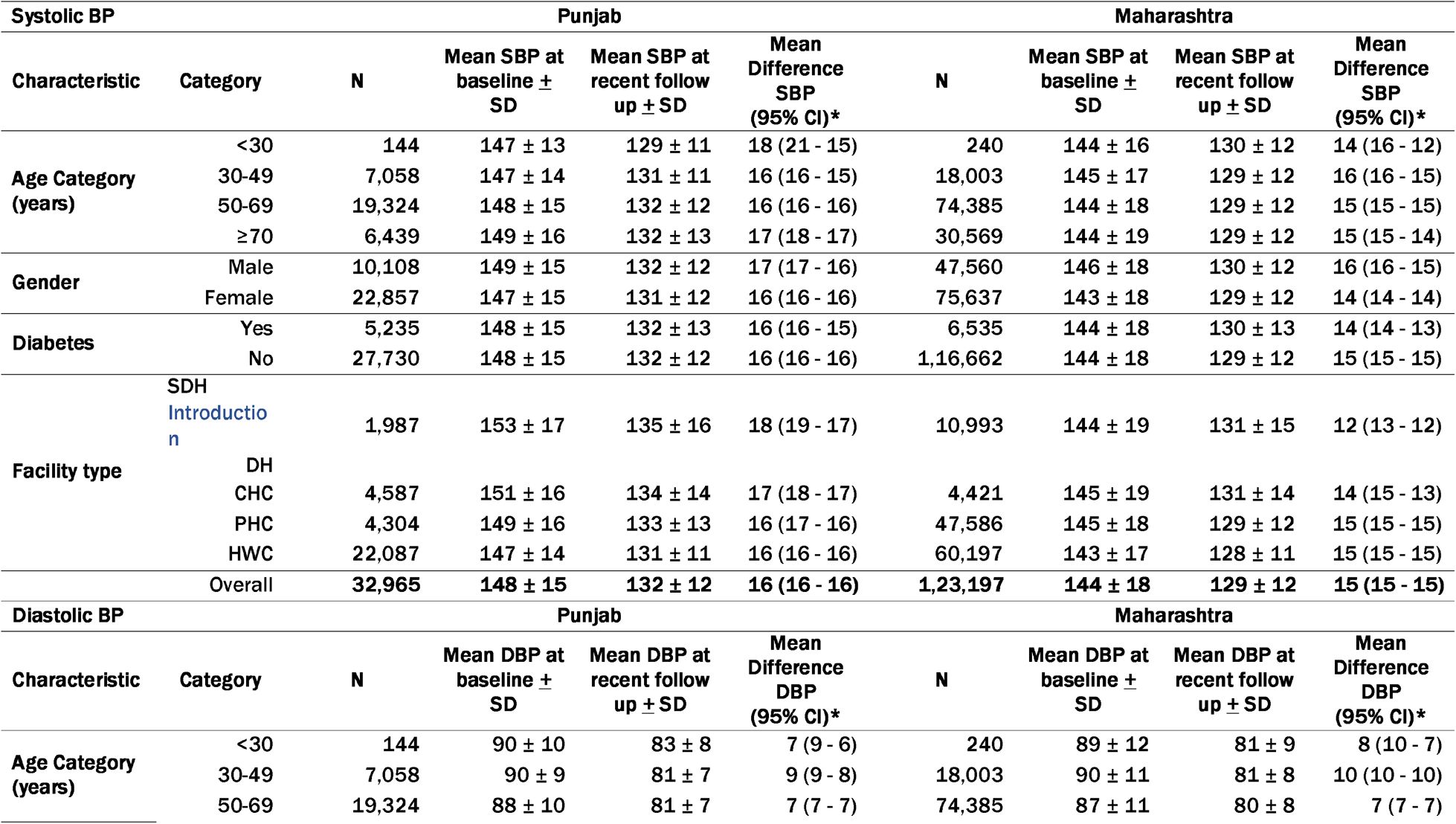
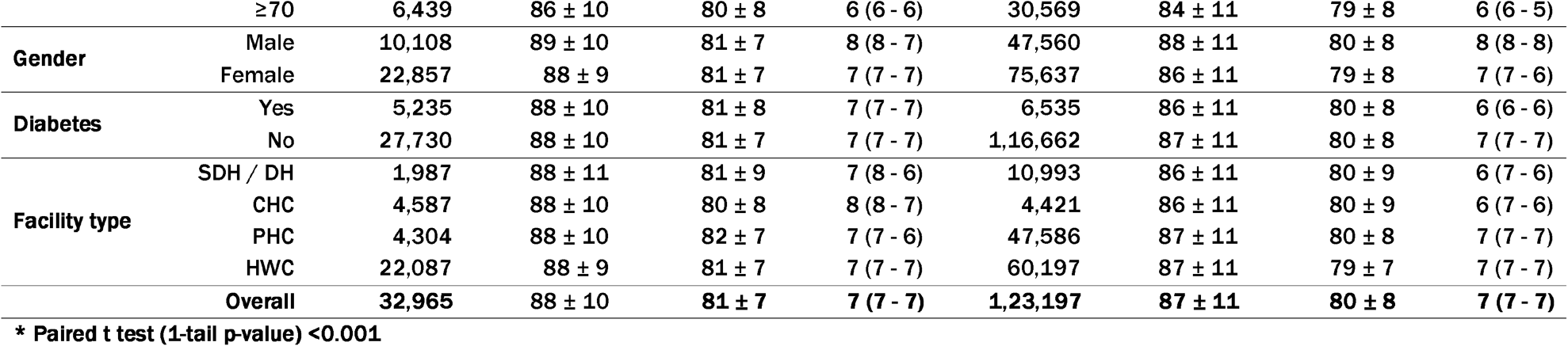
Baseline and follow-up blood pressure among individuals started on Amlo 5 mg on registration up to Dec 2021 and had at least one follow-up visit between 01 Jan 2022 to 31 Mar 2022 (excluding missing BP values at follow-up visit)

The mean (SD) diastolic blood pressure (DBP) decreased by 7.1 mmHg (p-value <0.001) in Punjab from 88.0 (9.5) at the baseline to 80.8 (7.4) during the recent visit (Table 2). In Maharashtra, the decline was 6.9 mmHg from 86.5 (11.0) to 79.6 (7.9) mmHg (p-value <0.001). Like SBP, the DBP also significantly declined from the baseline to the recent visit irrespective of age group, gender, presence of diabetes, facility type, and the current anti-hypertensive drug combination, ranging from 5.9 to 8.7 mmHg in Punjab and 5.5 to 9.6 mmHg in Maharashtra.

Among all people who were started with Amlodipine 5mg during registration and who were on follow-up with one of the IHCI protocol regimens, 70% in Punjab and 76% in Maharashtra had controlled BP only with amlodipine 5 mg (Table 3). In Punjab, at the second step with amlodipine 10 mg, the cumulative proportion with controlled BP increased by five percentage points to 75%. In the third step, with the combination of amlodipine 10 mg and telmisartan 40 mg, there was an additional increase of two percentage points to 77%. The proportion with controlled BP plateaued at 77% from step four of the protocol (i.e., the combination of amlodipine 10 mg and telmisartan 80 mg). We observed a similar pattern in Maharashtra. The proportion with controlled BP increased by five percentage points from 76% to 81% at the second step (combination of amlodipine 5 mg and telmisartan 40 mg) of the treatment protocol and plateaued at 82% from step three of the protocol (i.e., the combination of amlodipine 5 mg and telmisartan 80 mg).

**Table 3.** Blood pressure control by each step of the IHCI regimen among individuals started on Amlo 5 mg between 01 Jan 2018 and Dec 2021 and continuing treatment as per IHCI protocol regimens during follow-up, Jan-March, 2022.

| **Punjab** |  |  |  |  |
| --- | --- | --- | --- | --- |
| **Drug prescribed during second last visit** | **Follow up visit during Jan-March 2022** | **BP control at the recent visit during Jan-March 2022** |  | **Cumulative BP control among all those who had recent follow-up visit** |
|  | **N** | **n** | **%** |  |
| Amlo 5 | 29843 | 23292 | 78 | 70% |
| Amlo 10 | 2617 | 1796 | 69 | 75% |
| Amlo 10 +Telm 40 | 804 | 526 | 65 | 77% |
| Amlo 10 +Telm 80 | 133 | 77 | 58 | 77% |
| Amlo 10 +Telm 80 + Chlo 12.5 | 43 | 22 | 51 | 77% |
| Amlo 10 +Telm 80 + Chlo 25 | 10 | 4 | 40 | 77% |
|  | 33,450 | 25,717 | 77 | 77% |
| **Maharashtra** |  |  |  |  |
| **Drug prescribed during second last visit** | **Follow up visit during Jan-March 2022** | **BP control at the recent visit during Jan-March 2022** |  | **Cumulative BP control among all those who had recent follow-up visit** |
|  | **N** | **n** | **%** |  |
| Amlo 5 | 116376 | 96064 | 83 | 76% |
| Amlo 5 +Telm 40 | 8492 | 6153 | 72 | 81% |
| Amlo 5 + Telm 80 | 704 | 449 | 64 | 82% |
| Amlo 10 + Telm 80 | 214 | 131 | 61 | 82% |
| Amlo 10 +Telm 80 + Chlo 6.25 | 55 | 38 | 69 | 82% |
| Amlo 10 +Telm 80 + Chlo 12.5 | 1 | 0 | 0 | 82% |
|  | 1,25,842 | 1,02,835 | 82 | 82% |

Amlo - Amlodipine; Telm - Telmisartan; Chlo - Chlorthalidone

During registration, 17% in Punjab and 32% in Maharashtra had controlled BP. Among them, 87% in Punjab and 89% in Maharashtra continued to have controlled BP during follow-up. The proportion with controlled BP at the first step of the treatment protocol during follow-up was 71% and 76% for people with Grade I hypertension during registration in Punjab and Maharashtra, respectively (Table 4). Among people with Grade II hypertension during registration, 59% in Punjab and 65% in Maharashtra had controlled BP at the first step.

**Table 4.**
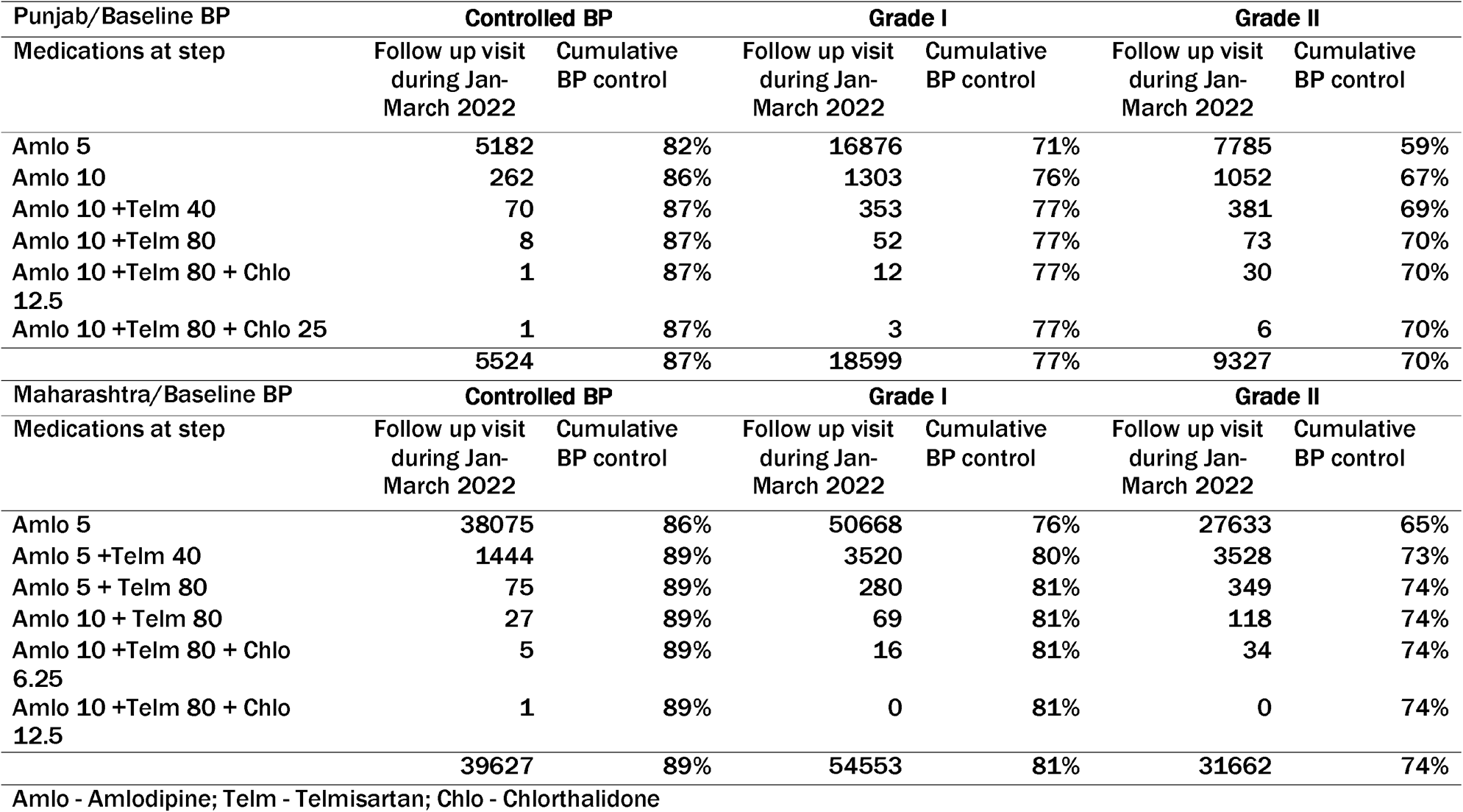
Blood pressure control by each protocol step & baseline BP grade among individuals started on Amlo 5 mg between 01 Jan 2018 and 31 Dec 2021 and was taking one of the IHCI protocol regimens during follow-up.

| **Punjab/Baseline BP** | **Controlled BP** |  | **Grade I** |  | **Grade II** |  |
| --- | --- | --- | --- | --- | --- | --- |
| **Medications at step** | **Follow up visit during Jan-March 2022** | **Cumulative BP control** | **Follow up visit during Jan-March 2022** | **Cumulative BP control** | **Follow up visit during Jan-March 2022** | **Cumulative BP control** |
| Amlol 5 | 5182 | 82% | 16876 | 71% | 7785 | 59% |
| Amlol 10 | 262 | 86% | 1303 | 76% | 1052 | 67% |
| Amlol 10 +Telm 40 | 70 | 87% | 353 | 77% | 381 | 69% |
| Amlol 10 +Telm 80 | 8 | 87% | 52 | 77% | 73 | 70% |
| Amlol 10 +Telm 80 + Chlo 12.5 | 1 | 87% | 12 | 77% | 30 | 70% |
| Amlol 10 +Telm 80 + Chlo 25 | 1 | 87% | 3 | 77% | 6 | 70% |
|  | 5524 | 87% | 18599 | 77% | 9327 | 70% |
| **Maharashtra/Baseline BP** | **Controlled BP** |  | **Grade I** |  | **Grade II** |  |
| **Medications at step** | **Follow up visit during Jan-March 2022** | **Cumulative BP control** | **Follow up visit during Jan-March 2022** | **Cumulative BP control** | **Follow up visit during Jan-March 2022** | **Cumulative BP control** |
| Amlol 5 | 38075 | 86% | 50668 | 76% | 27633 | 65% |
| Amlol 5 +Telm 40 | 1444 | 89% | 3520 | 80% | 3528 | 73% |
| Amlol 5 + Telm 80 | 75 | 89% | 280 | 81% | 349 | 74% |
| Amlol 10 + Telm 80 | 27 | 89% | 69 | 81% | 118 | 74% |
| Amlol 10 +Telm 80 + Chlo 6.25 | 5 | 89% | 16 | 81% | 34 | 74% |
| Amlol 10 +Telm 80 + Chlo 12.5 | 1 | 89% | 0 | 81% | 0 | 74% |
|  | 39627 | 89% | 54553 | 81% | 31662 | 74% |

Amlol - Amlodipine; Telm - Telmisartan; Chlo - Chlorthalidone

Adding Telmisartan 40 mg (in Maharashtra) or Amlodipine 10 mg (in Punjab) in the second step increased the cumulative control during follow-up by four to five percentage points among those with Grade I BP during registration. The individuals with Grade II baseline BP had an eight percentage points increase in cumulative BP control at the third step in both states. The cumulative control reached a plateau from the third or fourth step across all grades.

## Discussion

We demonstrated the effectiveness of simple drug- and dose-specific treatment protocols in public sector primary and secondary care health facilities at scale in two Indian states. There is ample evidence that regular treatment with anti-hypertensive drugs controls blood pressure. However, the scalability of hypertension programs requires protocols to enable efficient procurement and rapid training of many healthcare workers. Our results are based on real-time data in a programmatic setting in a large cohort of people with hypertension. Although the protocols and health systems differed in the two states, effectiveness was similar. Both states implemented other IHCI strategies to strengthen the health system, making the program comparable in training, supervision, and information system. The project team worked closely with the state public health departments to streamline the procurement and distribution of drugs within the existing health system. The drugs were given free of cost at all the clinics, ensuring affordability was not a barrier to care.

The effectiveness of simple treatment protocols in primary care can be an enabler in improving treatment coverage in India and other LMICs. Treatment of hypertension is one of the best buys to reduce cardiovascular mortality, per the WHO guidelines [13]. Globally, 38% of men and 47% of women were taking treatment for hypertension in 2019 [14]. Physicians often tend to have their drug preferences. However, it is not feasible to scale hypertension programs in low-resource settings unless there is a reliable supply of a few drugs free of cost to the patient. Despite robust evidence from clinical trials and programs in high-income countries regarding the benefits of hypertension control to reduce CVD mortality, treatment coverage, and blood pressure control are inadequate. The WHO-HEARTS package launched in 2017 recommended simple drug and dose-specific protocols [9]. The World Health Organization (WHO) recently published revised hypertension management guidelines [3]. The IHCI project in India developed locally relevant protocols based on the HEARTS package. Several countries have recently developed drug and dose-specific protocols to accelerate hypertension control [15]. A study from 12 countries in the Americas reported involvement of ministries and experts facilitated the uptake of protocols. We finalized the protocols in consultation with various stakeholders within each state, which was an enabler in the acceptance of protocols not only in the project districts but also state-wide. In addition to clinical evidence, the ability to procure the drugs at scale within available budgets was considered while deciding protocols. The high BP control in the study among people who continued the treatment is very encouraging. One major factor influencing overall program effectiveness was the lack of regular follow-up for a sub-group of people enrolled for treatment in the study quarter. This reinforces the importance of retention in care in addition to protocol-based treatment and the availability of free drugs. We also observed that a fraction of people with BP control had uncontrolled BP at follow-up, emphasizing the importance of continuous treatment and follow-up even after control is achieved.

We reported BP control at the various steps of the protocol using the cut-off SBP 140 mmHg and 90 mmHg. In addition, the average SBP decline of 15-16 mmHg was observed in two states. Protocol-based treatment will benefit all patients whose BP decreased, even if they did not achieve BP control. We could have overestimated the decline due to regression to mean, a well-documented phenomenon in hypertension randomized trials [16].

A meta-analysis of randomized trials showed a 10% reduction in cardiovascular events for every five mmHg reduction in the SBP [17]. The average SBP decline was lower in a multicounty RCT from Bangladesh, Pakistan, and Sri Lanka, where primary care multicomponent intervention was implemented. The trial included 2645 adults with hypertension and reported a decline of 9.0 mm Hg in the intervention group compared to 3.9 mm Hg in the control group [18]. Regression to mean might account for the partial decline in blood pressure. Even after discounting the partial decline to regression to mean and considering the evidence from other settings, the BP change was sufficient to reduce CVD risk substantially.

The BP control and average decline were similar in the two states despite using two different protocols. In Punjab, Amlodipine was escalated from 5 mg to 10 mg in the second step. On the other hand, Telmisartan was added at the second step in the Maharashtra protocol. The increase in control at the second step was similar irrespective of the protocol. This observation has implications for hypertension programs in low-resource settings. Amlodipine is one of the least expensive antihypertensives which can be procured at scale with limited budgets in the NCD programs. Protocols with Amlodipine for the first and second steps might be more feasible and scalable. The selection of a second drug should be based on availability, cost, and local context. In the two states, the difference in cost between the regimens was substantial: Using Amlodipine in the first two steps reduced the cost by about 20% (2.05 USD vs. 2.50 USD) per patient treated per year [19].

Some guidelines recommend a Single pill fixed-dose combination (FDC) as a first-line drug [3,4]. Our findings suggest that FDC as the first-line drug for all individuals with hypertension might not be necessary for large-scale programs in low-resource settings. Rather, an uninterrupted supply of a low-cost drug is a more important facilitator for scaling hypertension programs. India has over 200 million people with hypertension; the treatment coverage is only 14.5% [1]. FDC for all, as the first line, will increase the cost and may not be scalable for large populations. We need further research to assess if the FDC as a first-line drug-based protocol will help improve BP control among people with SBP above 160 mmHg and DBP above 100 mmHg at the diagnosis.

Our study has several limitations. The study’s major limitation was that we could only include individuals already on treatment with Amlodipine 5 mg or newly diagnosed and initiated on Amlodipine 5 mg, which is the first step of the protocol. Patients already on treatment on other drugs or started on non-protocol drugs due to physician preference could not be included. The second limitation was the inability to document the blood pressure of people who did not return to the clinic. We compared the characteristics of the two groups – included and excluded from the analysis to understand if they differed. The major differences were a higher proportion of diabetes (32% vs. 8%), a lower proportion of Grade I hypertension at baseline (36% vs. 46%), and a higher proportion of controlled BP at baseline (28% vs. 32%) among those excluded than included. The third limitation was the inability to understand the protocol’s effectiveness beyond the second step, as the number who progressed to that stage was relatively small. The fourth limitation was that we could not document the side effects, which might have led to loss-to-follow-up or drug changes, not as per protocol. Based on the unpublished data from a preliminary survey conducted in seven sentinel clinics under the IHCI project, about 12% of the individuals on Amlodipine 10mg had pedal edema. Drug shortage and physician preferences might also have led to deviations from the protocol. As the data was collected in programmatic conditions, it was not feasible to document the information at scale. However, we recommend collecting the data on side effects in a sample of clinics hereafter to overcome the limitation. The fifth limitation was a possible overestimation of the effectiveness due to regression to mean effect [16]. However, there are many strengths in our study. The study had a large sample size and real-time patient tracking using a real-time app-based information system in the programmatic setting. Nurses recorded BP readings and medications as and when patients visited the clinic. There was no data transfer from paper to digital, which minimized the errors. We could verify when BP was recorded and the time to record the visit. Tracking the patterns enabled the identification of any major deviations in any of the clinics. In such situations, the supervisor conducted a supportive supervision visit and addressed if there were any training gaps or any other challenges.

We demonstrated that simple treatment protocols effectively achieve blood pressure control at scale in the real-world setting under programmatic conditions. Despite programmatic challenges, many people who continued treatment achieved blood pressure control with one or two low-cost drugs. Our study findings have relevance for other Indian states and similar LMIC settings. We recommend treatment protocols starting with a single low-cost drug and escalating with the same or another antihypertensive drug subject to availability in low-resource settings. Simple treatment protocols can facilitate the rapid scaling of hypertension treatment to large populations in primary care globally. We need further research to understand the effectiveness of protocols which include fixed-dose combination at the second step and initiation of combination drug as the first step for people with SBP above 160 mmHg and DBP above 100 mmHg. Future studies may also document the side effects of anti-hypertensive drugs and the reasons for loss-to-follow-up.

## Data Availability

Data are available upon reasonable request

## Acknowledgement

We thank the “Resolve to Save Lives” team for their valuable technical inputs in the design and implementation of the “India Hypertension Control Initiative” project. We thank doctors, nurses, and all the health officials in various project districts for their support in the project implementation. We thank Senior Treatment Supervisors for their role in capacity building and supportive supervision in the project districts.

## Authors’ contributions

PK was involved in conceptualization, design, methodology and supervision of the IHCI project, and data analysis, preparation of the first draft, and revision of the current manuscript. MS and VV were part of the data analysis, interpretation, preparation of the first draft, and manuscript revision. PJ and SSG played a lead role in implementing the intervention in the respective states. They were also involved in the supervision, data interpretation, critical review, and manuscript revision. AK, KD, PG, SKS, AK, and PP contributed to methodology, supervision, data acquisition, and manuscript revision. MS, AKP, RS, and BB were part of conceptualization, design, methodology, critical review and manuscript revision. AW, NK, VB, BD, TC, SK, LS, and SDB were involved in implementing the interventions at the district or state level, supervision, data acquisition and data management, critical review, and manuscript revision. PK and MS contributed equally to this paper and designated as co-first authors. All authors have read and agreed to the contents of the manuscript.

## Data sharing statement

Data are available upon reasonable request

---

**Supplementary Table 1.**
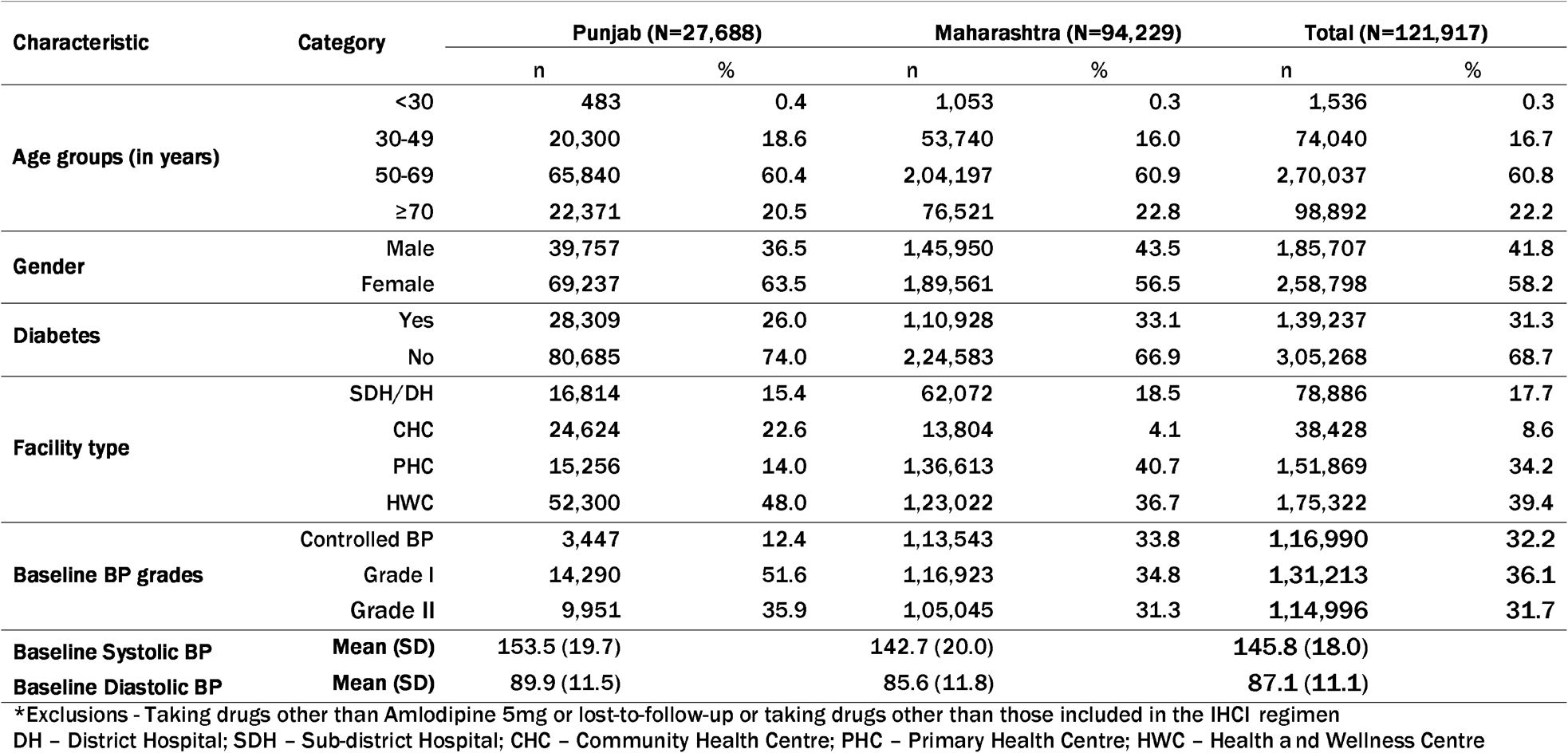
Characteristics of individuals registered under IHCI but excluded* from the analysis.

**Supplementary Table 2.**
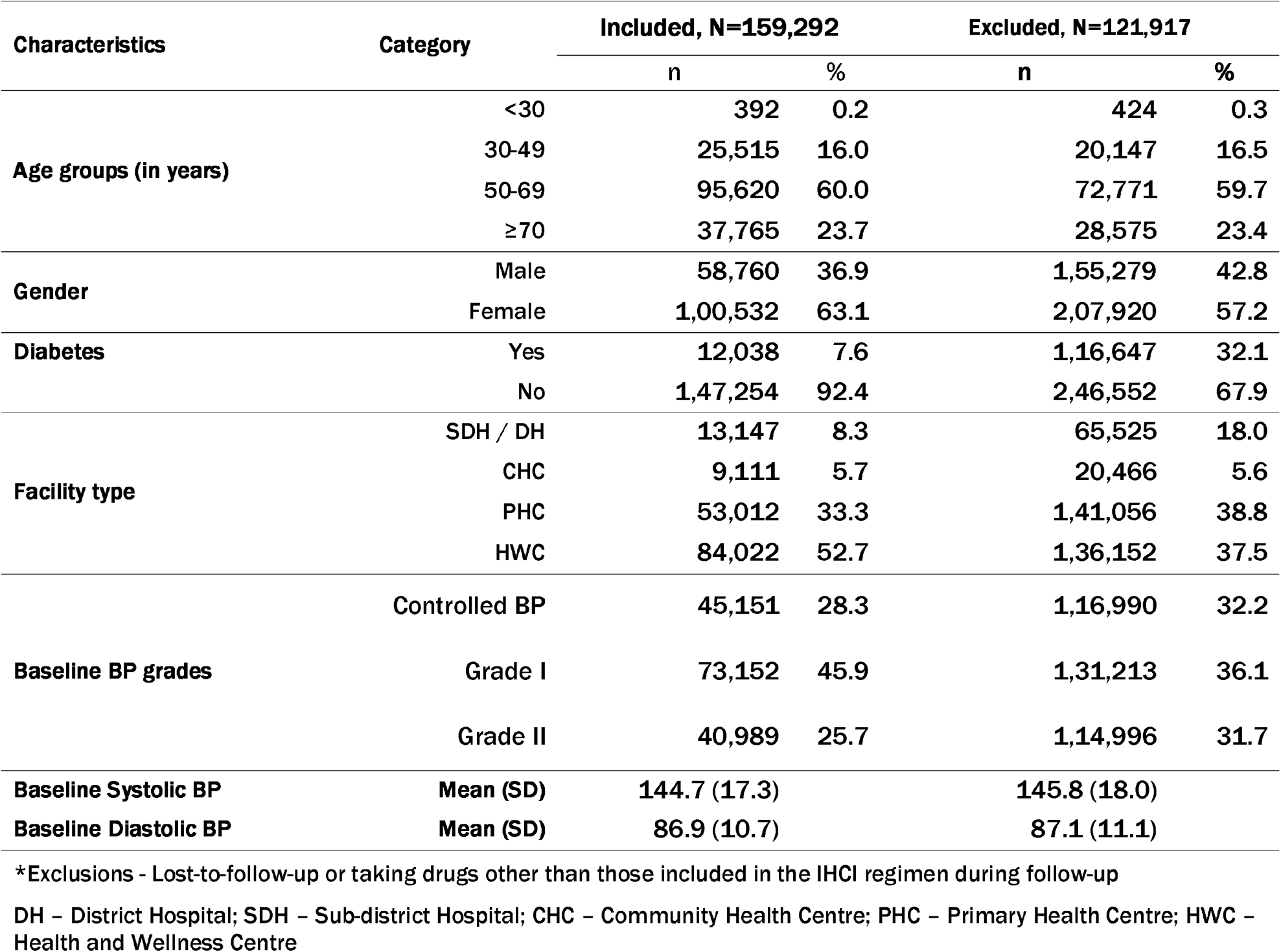
Comparison of characteristics of individuals included & excluded* from the analysis.

**Supplementary Figure 1.**
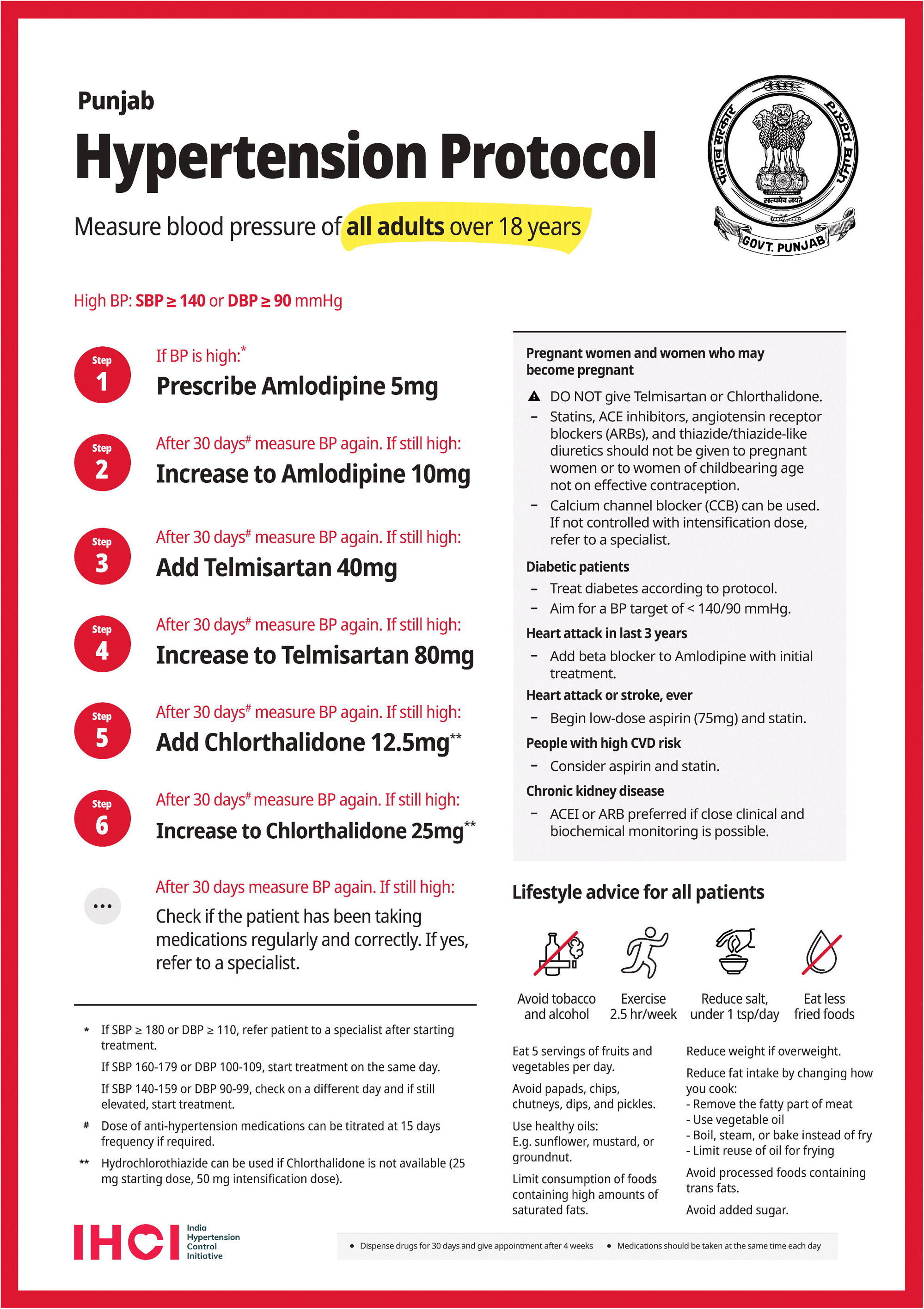
Hypertension treatment protocol followed in IHCI districts, Punjab, India

**Supplementary Figure 2.**
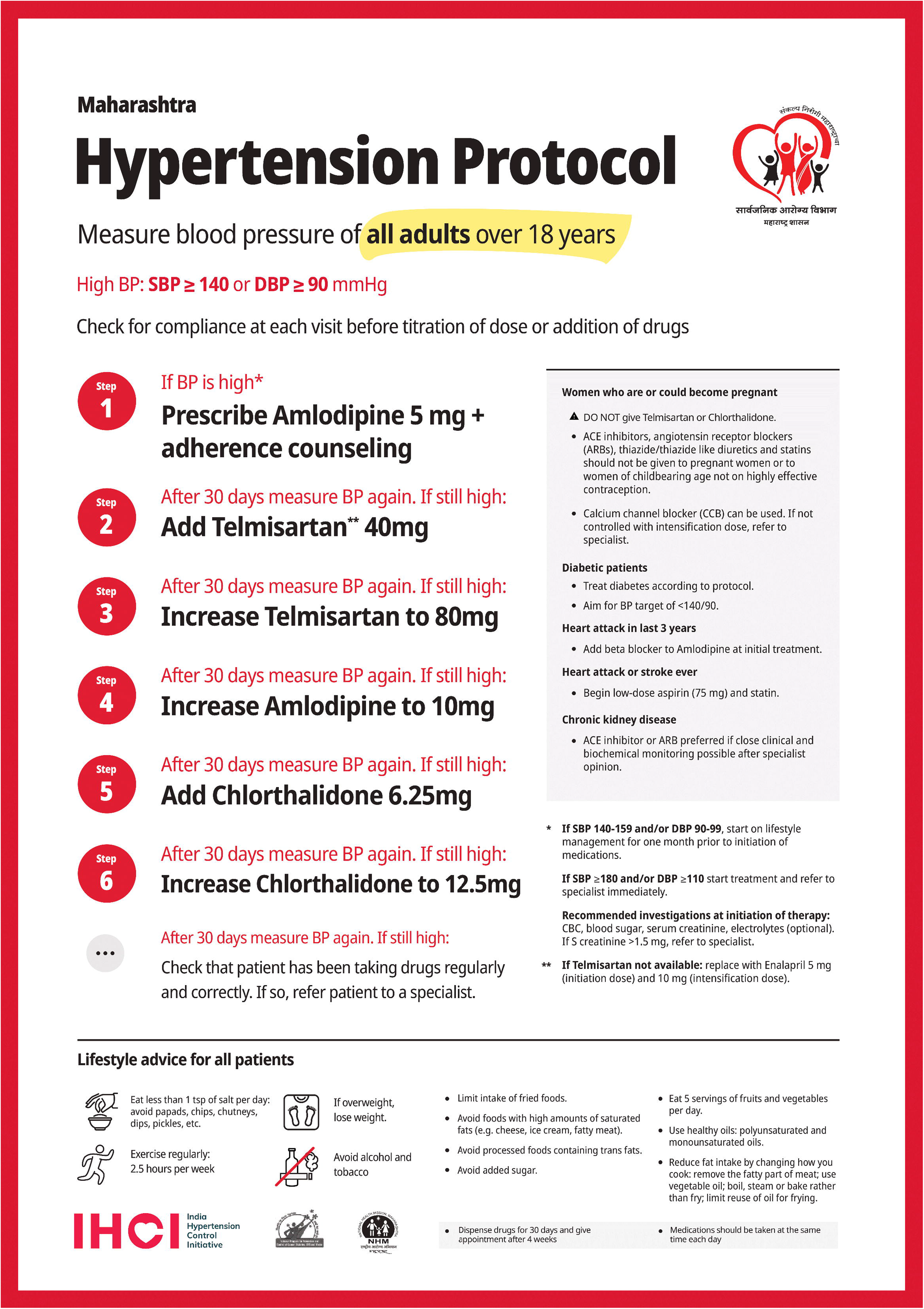
Hypertension treatment protocol followed in IHCI districts, Maharashtra, India

