## Supplementary Table 1 for "India Hypertension Control Initiative- Blood Pressure Control using Drug and Dose-Specific Standard Treatment Protocol at Scale in Punjab and Maharashtra, India, 2022"

**Supplementary Table 1. Characteristics of individuals registered under IHCI but excluded* from the analysis**

| **Characteristic** | **Category** | **Punjab (N=27,688)** | | | **Maharashtra (N=94,229)** | | | **Total (N=121,917)** | |
| --- | --- | --- | --- | --- | --- | --- | --- | --- | --- |
|  |  | n | % | n | | % | n | | % |
| **Age groups (in years)** | <30 | 483 | 0.4 | 1,053 | | 0.3 | 1,536 | | 0.3 |
|  | 30-49 | 20,300 | 18.6 | 53,740 | | 16.0 | 74,040 | | 16.7 |
|  | 50-69 | 65,840 | 60.4 | 2,04,197 | | 60.9 | 2,70,037 | | 60.8 |
|  | ≥70 | 22,371 | 20.5 | 76,521 | | 22.8 | 98,892 | | 22.2 |
| **Gender** | Male | 39,757 | 36.5 | 1,45,950 | | 43.5 | 1,85,707 | | 41.8 |
|  | Female | 69,237 | 63.5 | 1,89,561 | | 56.5 | 2,58,798 | | 58.2 |
| **Diabetes** | Yes | 28,309 | 26.0 | 1,10,928 | | 33.1 | 1,39,237 | | 31.3 |
|  | No | 80,685 | 74.0 | 2,24,583 | | 66.9 | 3,05,268 | | 68.7 |
| **Facility type** | SDH/DH | 16,814 | 15.4 | 62,072 | | 18.5 | 78,886 | | 17.7 |
|  | CHC | 24,624 | 22.6 | 13,804 | | 4.1 | 38,428 | | 8.6 |
|  | PHC | 15,256 | 14.0 | 1,36,613 | | 40.7 | 1,51,869 | | 34.2 |
|  | HWC | 52,300 | 48.0 | 1,23,022 | | 36.7 | 1,75,322 | | 39.4 |
| **Baseline BP grades** | Controlled BP | 3,447 | 12.4 | 1,13,543 | | 33.8 | 1,16,990 | | 32.2 |
|  | Grade I | 14,290 | 51.6 | 1,16,923 | | 34.8 | 1,31,213 | | 36.1 |
|  | Grade II | 9,951 | 35.9 | 1,05,045 | | 31.3 | 1,14,996 | | 31.7 |
| **Baseline Systolic BP** | **Mean (SD)** | 153.5 (19.7) |  | 142.7 (20.0) | |  | 145.8 (18.0) | |  |
| **Baseline Diastolic BP** | **Mean (SD)** | 89.9 (11.5) |  | 85.6 (11.8) | |  | 87.1 (11.1) | |  |
| *Exclusions - Taking drugs other than Amlodipine 5mg or lost-to-follow-up or taking drugs other than those included in the IHCI regimen  DH – District Hospital; SDH – Sub-district Hospital; CHC – Community Health Centre; PHC – Primary Health Centre; HWC – Health and Wellness Centre | | | | | | | | | |
