## Supplementary Table 2 for "India Hypertension Control Initiative- Blood Pressure Control using Drug and Dose-Specific Standard Treatment Protocol at Scale in Punjab and Maharashtra, India, 2022"

**Supplementary Table 2. Comparison of characteristics of individuals included & excluded* from the analysis**

| **Characteristics** | **Category** | **Included, N=159,292** | | **Excluded, N=121,917** | |
| --- | --- | --- | --- | --- | --- |
|  |  | n | % | **n** | **%** |
| **Age groups (in years)** | <30 | 392 | 0.2 | 424 | 0.3 |
|  | 30-49 | 25,515 | 16.0 | 20,147 | 16.5 |
|  | 50-69 | 95,620 | 60.0 | 72,771 | 59.7 |
|  | ≥70 | 37,765 | 23.7 | 28,575 | 23.4 |
| **Gender** | Male | 58,760 | 36.9 | 1,55,279 | 42.8 |
|  | Female | 1,00,532 | 63.1 | 2,07,920 | 57.2 |
| **Diabetes** | Yes | 12,038 | 7.6 | 1,16,647 | 32.1 |
|  | No | 1,47,254 | 92.4 | 2,46,552 | 67.9 |
| **Facility type** | SDH / DH | 13,147 | 8.3 | 65,525 | 18.0 |
|  | CHC | 9,111 | 5.7 | 20,466 | 5.6 |
|  | PHC | 53,012 | 33.3 | 1,41,056 | 38.8 |
|  | HWC | 84,022 | 52.7 | 1,36,152 | 37.5 |
| **Baseline BP grades** | Controlled BP | 45,151 | 28.3 | 1,16,990 | 32.2 |
|  | Grade I | 73,152 | 45.9 | 1,31,213 | 36.1 |
|  | Grade II | 40,989 | 25.7 | 1,14,996 | 31.7 |
| **Baseline Systolic BP** | **Mean (SD)** | 144.7 (17.3) |  | 145.8 (18.0) |  |
| **Baseline Diastolic BP** | **Mean (SD)** | 86.9 (10.7) |  | 87.1 (11.1) |  |
| *Exclusions - Lost-to-follow-up or taking drugs other than those included in the IHCI regimen during follow-up | | | | | |
| DH – District Hospital; SDH – Sub-district Hospital; CHC – Community Health Centre; PHC – Primary Health Centre; HWC – Health and Wellness Centre | | | | | |
